# Mother-Centric Motivational Interviewing for Behavioral Change and Obesity Prevention in Preschool-Aged Children in Bangladesh: A Pilot Study

**DOI:** 10.1101/2025.06.23.25330164

**Authors:** Mohammad Sorowar Hossain, Farhin Islam, Shameema Ferdous, Anthony D Okely

## Abstract

**Introduction:** Childhood obesity is a rising public health concern, particularly in low- and middle-income countries (LMICs) like Bangladesh, where the rate of obesity in children has increased. South Asian countries, traditionally burdened by undernutrition, now face a growing challenge of obesity. This study aimed to evaluate the impact of a mother-centric Motivational Interviewing (MI) intervention on childhood obesity prevention and behavioral outcomes in preschool-aged children in Bangladesh.

**Methods:** A quasi-experimental, pre-post intervention study was conducted in Jamalpur district town between June 2023 and January 2024. The intervention comprised two school-based MI counseling sessions for mothers, while the control group received a general health education session except for obesity. Behavioral and anthropometric data were collected at baseline and after eight months. The primary analysis employed Difference-in-Differences (DID) regression to estimate intervention effects, using ordered probit and OLS models for categorical and continuous outcomes, respectively.

**Results:** The intervention significantly improved physical activity frequency (β = 0.661, p < 0.05), sleep quality (β = 1.090, p < 0.05), and reduced mothers screen time (β = –1.102, p < 0.10). No significant changes were observed in dietary behaviors, childrens screen use, or anthropometric indicators (WAZ, HAZ, BAZ). Parallel trend violations in some variables may limit causal inference.

**Conclusion:** The MI-based intervention led to positive behavioral changes, particularly in physical activity and sleep quality, among children and mothers. However, it did not result in significant improvements in anthropometric outcomes, likely due to the short intervention period, high dropout rate, and small sample size. This study suggests that MI interventions, when culturally adapted, hold promise for promoting healthier lifestyles in LMICs, but further research with longer follow-up is needed to observe lasting impacts on obesity prevention.

## Introduction

Childhood obesity is recognized globally as one of the most pressing public health challenges of the 21st century[1]. Over two-thirds of children with obesity or being overweight live in low- and middle-income countries, where the rate of childhood obesity has risen significantly in recent years[2].

South Asia, traditionally burdened by undernutrition, is now facing a growing challenge of childhood obesity. A systematic review of 152 studies conducted between 1994 and 2023 reported pooled prevalence rates of 6.6% for obesity, 12.4% for overweight, and 19.3% for combined overweight and obesity among children and adolescents in South Asia. Notably, Bangladesh showed a higher prevalence of overweight at 13.6%, highlighting an emerging public health concern [3].

In Bangladesh, our community level study found that 14% of preschool-aged children were overweight or obese, but mothers perceived only about 3% of their children as overweight or obese[4]. The same study revealed that childhood obesity was not well understood by primary caregivers (mothers), who often thought it as a sign of good health. Nearly 65% of mothers did not view childhood obesity as a health concern, and over two-thirds (∼69%) were unaware of its potential health consequences[4].

Childhood obesity is influenced by many lifestyle, socioeconomic, genetic, and environmental factors[5]. Systematic reviews have consistently shown that physical activity (PA) is inversely associated with adiposity in children, indicating that higher levels of PA are linked to lower body fat [6–8]. More recently, research has emphasized the importance of 24-hour movement behaviors (physical activity, screen time, and sleep duration) is significantly associated with childhood obesity [9]. The World Health Organization’s Commission on Ending Childhood Obesity has also highlighted the role of these movement behaviors in obesity prevention among children[10].

Preventing overweight and obesity) early in life is crucial, as children with excess weight are more likely to become obese adults[11]. Targeting interventions during the preschool years may be particularly effective, as this period is critical for the development of long-term dietary and physical activity habits[12].

Research has shown that unhealthy weight gain is preventable if addressed early by promoting healthy behaviors. Parents play a pivotal role in shaping children’s healthy behaviors, including diet, physical activity, sleep, and screen time. Factors such as poor feeding practices, indulgent parenting, high parental stress, and an unsupportive home environment have been linked to increased risk of childhood obesity [13]. Thus, encouraging and supporting the development of healthy habits within the family, along with equipping parents with the knowledge and resources to foster these behaviors, is essential.

Motivational Interviewing (MI) is a evidence based, goal-oriented counseling method that aims to encourage individuals overcome ambivalence and enhance their internal motivation to change. It is particularly effective in promoting behavior change related to health, addiction, and lifestyle choices[14]. Systematic reviews reveals that motivational interviewing (MI) interventions have been associated increased levels of physical activity, reduced screen time, and lower intake of sugary foods and beverages[15]. Evidence suggests that MI can significantly promote weight loss in children with obesity[16].

To the best of our knowledge, no intervention studies aiming on preventing unhealthy weights have been conducted in Bangladesh. In this country, most mothers stay home and care for their children[4]. South Asian mothers, in particular, dedicate their lives to their children’s well-being and are deeply concerned about their academic career[17]. Intervention programs, therefore, targeting mothers would likely be cost-effective, especially if they focus on educating mothers about the risk factors of unhealthy weights.

This study aimed (i) to assess the impact of a mother-centric, MI-based intervention on reducing overweight/obesity and improving behavioral outcomes (e.g., increased physical activity, reduced screen time, improved sleep duration and quality, and reduced consumption of sugary foods and drinks) among children in the early years and (ii) to evaluate the strengths and weaknesses of the mother-centric MI approach in the community setting of Bangladesh.

## Methods

### Study Design and Setting

This study was a quasi-experimental, community-based pilot intervention conducted in Jamalpur Paurashava, a urban municipality in northern Bangladesh. The study followed a pre-post design with a non-randomised comparison (control) group and was implemented from June 2023 to January 2024. The objective was to evaluate the impact of a mother-centric behavioural counselling strategy—rooted in Motivational Interviewing (MI)—on childhood obesity prevention and related lifestyle behaviours. Data were collected at two time points: baseline (pre-intervention) and eight months after the intervention (post-intervention). The intervention targeted four primary behavioural domains: physical activity, screen time, sleep hygiene, and dietary habits. Mothers in the control group participated in a single-session general health workshop unrelated to nutrition or obesity.

### Participant Recruitment and Selection

Participants were mother-child dyads recruited from four purposively selected schools. These schools were chosen from a pool of twelve schools that had participated in an earlier cross-sectional survey conducted by the research team, which examined maternal perceptions of childhood obesity[4]. School selection was guided by logistical feasibility and institutional willingness to collaborate.

Children aged 4 to 7 years, enrolled in preschool or kindergarten classes, were eligible to participate. Mothers were required to provide written informed consent. Dyads were excluded if the child or parent had any serious physical or psychological condition that could compromise participation. Recruitment was facilitated by school administrators who distributed study invitations to eligible families. Two schools were assigned to the intervention group and two to the control group. Assignment was not randomised due to logistical constraints, including concerns about cross-group contamination and differing school schedules.

Initially we targeted 260 mother-child pairs (130 for treatment and 130 for control group) in this study. However, at baseline, 118 mothers were enrolled in the intervention group and 104 in the control group. At post-intervention follow-up, data were collected from 60 mother-child pairs in the intervention group and 45 in the control group, reflecting dropout rates of 49.1% and 56.7%, respectively (Figure 1). This high drop-out is mainly due to school session transitions, with several children transferring to other institutions. The relatively high attrition was likely influenced by overlapping school session transitions, with many families relocating their children to other institutions.

**Figure 1:**
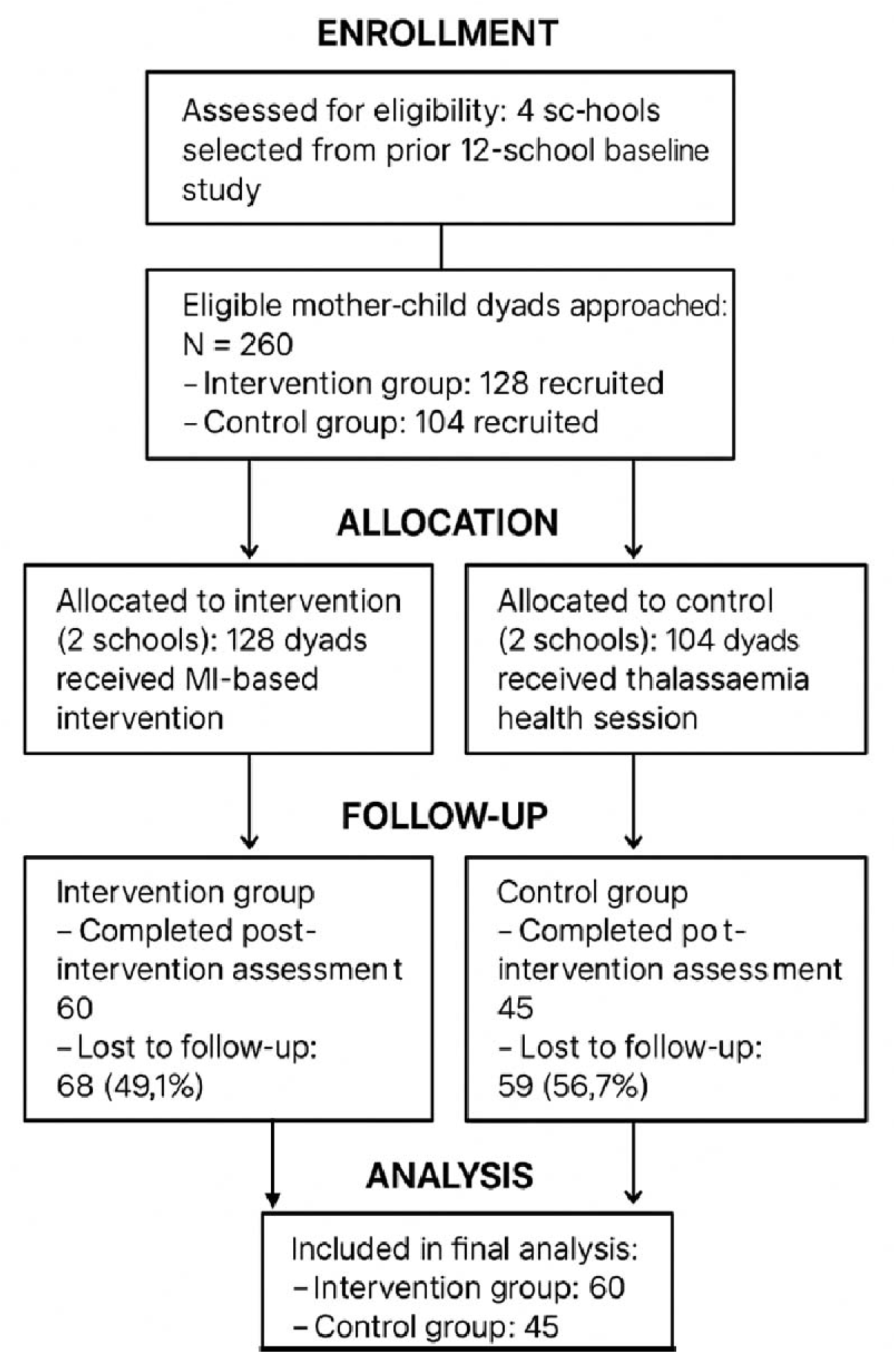
Participant flow

### Intervention Design

The intervention was based on the principles of Motivational Interviewing (MI), a client-centred, goal-oriented counselling approach that facilitates health behaviour change by enhancing intrinsic motivation and resolving ambivalence. The aim was to empower mothers to modify home-based routines that influence their children’s health behaviours. The intervention included two in-person group counselling sessions, each lasting approximately two hours and spaced over a four-month period. The sessions were delivered on school premises by the principal investigator, who has experienced in MI related public health communication.

The counselling covered key areas such as understanding the health consequences of childhood obesity, strategies to encourage regular physical activity and improve sleep hygiene, techniques to manage and reduce screen time for both mothers and children, and approaches to improve family dietary habits, including limiting junk food and sugar-sweetened beverage consumption. To reinforce learning and behavioural change, mothers received educational handouts summarising the session content. They were also regularly provided with a “Healthy Lifestyle Monitoring Form,” distributed and collected through class teachers. This form allowed mothers to report and reflect on their child’s behaviours across four domains: physical activity, screen time, sleep, and dietary practices. The intervention was designed to align with school schedules and avoid disruption to academic activities.

### Control Group

The control group received a one-time general health education session that focused on thalassaemia prevention. Thalassaemia was selected as it is a recognised public health issue in Bangladesh but unrelated to lifestyle or obesity, thereby limiting the potential for contamination of outcomes. No follow-up contact or materials were provided to the control group participants.

### Implementation Fidelity and Monitoring

To ensure consistent implementation, a standardised guide was used for all counselling sessions. Completion of the biweekly monitoring forms was tracked by class teachers. Informal feedback was collected from participating mothers and school staff to assess the relevance and clarity of the intervention materials and delivery.

### Data Collection

Data were collected at baseline and post-intervention using structured questionnaires administered face-to-face by trained enumerators with backgrounds in public health and nutrition. Data collection was conducted in Bangla and overseen by field supervisor to ensure quality and consistency. The questionnaire captured sociodemographic information, including the age, education, occupation, and income of the parents, and the number of children in the household.

Child lifestyle behaviours were assessed through maternal reports including physical activity (number of days with at least one hour of activity per week), screen use patterns (such as use for teaching, calming, or distraction), dietary habits (junk food and sugary beverage consumption, use of screens during meals, and frequency of family meals), and sleep (average night-time sleep duration and subjective sleep quality, rated on a 0–7 scale). This questionnaire was adapted from the SUNRISE study (International study of 24-h movement behaviors of early years)[18]. Anthropometric data were also collected as per our previous study [19].

Children’s height and weight were measured using calibrated equipment following WHO guidelines. Z-scores for weight-for-age (WAZ), height-for-age (HAZ), and BMI-for-age (BAZ) were computed using WHO AnthroPlus software, following the 2007 growth reference standards. Each indicator was categorised using standard WHO classifications.

### Data Management and Analysis

Data were entered into a secure REDCap database and exported to STATA for analysis. All behavioural variables were converted into long format to facilitate pre-post comparisons. Descriptive statistics were calculated to summarise participant characteristics. Differences between treatment and control groups at baseline were assessed using chi-square tests for categorical variables and independent t-tests for continuous variables.

To estimate the intervention’s effect, Difference-in-Differences (DID) regression models were employed. These models calculated the average treatment effect by comparing changes in outcomes over time between the intervention and control groups. Ordered Probit regression was used for ordinal outcome variables such as sleep quality, frequency of physical activity, and dietary behaviours, while Ordinary Least Squares (OLS) regression was used for continuous outcomes such as sleep duration and screen time (measured in hours). Statistical significance was set at p < 0.05. Standard errors were clustered at the individual level. For outcomes with significant baseline differences (e.g., screen-related behaviours), the assumption of parallel trends may have been violated, and findings should be interpreted cautiously.

### Ethical Considerations

The study protocol was approved by the Institutional Review Board (IRB) of [Independent University, Bangladesh, Memo no: 2022-SLES-02 ]. All participants provided written informed consent prior to data collection. Confidentiality was maintained throughout, and participants were free to withdraw at any point without consequence.

## Results

### Participant Characteristics

Table 1 represents participant characteristic. A total of 105 preschool-aged children and their mothers participated in the study, with 60 assigned to the intervention group and 45 to the control group. The mean age of the children was 5.6 years (SD ± 0.71), with 60% being male. Most mothers were between 26 and 35 years old, and the majority had completed higher secondary education (39%) or were graduates (43%). Nearly 89% of mothers identified as housewives, and most lived with their husbands (90%). Fathers were generally well-educated, with about half holding graduate degrees, and were primarily employed in service (59%) or business (38%). Regarding income, nearly half of the families (47%) earned between 25,000 and 49,999 BDT per month. Most families (55%) had two children.

**Table 1:** Sociodemographic characteristics of participants.

| Variable | Characteristics | Freq. | Percent |
| --- | --- | --- | --- |
| Intervention group | Control | 45 | 42.86 |
|  | Treatment | 60 | 57.14 |
| Age of child (in years) | Mean ± SD | N = 105 | 5.6 ± 0.71 |
| Sex of child | Male | 63 | 60.00 |
|  | Female | 42 | 40.00 |
| Age of mother | 21-25 | 27 | 25.71 |
|  | 26-30 | 30 | 28.57 |
|  | 31-35 | 35 | 33.33 |
|  | >35 | 13 | 12.38 |
| Education of mother | Primary | 4 | 3.81 |
|  | Secondary | 15 | 14.29 |
|  | Higher Secondary | 41 | 39.05 |
|  | Graduate | 45 | 42.86 |
| Occupation of mother | Housewife | 93 | 88.57 |
|  | Service holder | 9 | 8.57 |
|  | Entrepreneur (Business) | 3 | 2.86 |
| Marital status | Living with husband | 95 | 90.48 |
|  | Living separately | 10 | 9.52 |
| Education of father | Primary | 6 | 5.71 |
|  | Secondary | 13 | 12.38 |
|  | Higher Secondary | 34 | 32.38 |
|  | Graduate | 52 | 49.52 |
| Occupation of father | Service holder | 62 | 59.05 |
|  | Businessman | 40 | 38.10 |
|  | Farmer | 1 | 0.95 |
|  | Others | 2 | 1.90 |
| Monthly family income | Less than 10000 | 12 | 11.43 |
|  | 10001-24999 | 27 | 25.71 |
|  | 25000-49999 | 49 | 46.67 |
|  | More than 50000 | 17 | 16.19 |
| Number of children in the family | 1 | 31 | 29.52 |
|  | 2 | 58 | 55.24 |
|  | 3 | 14 | 13.33 |
|  | >3 | 2 | 1.90 |

### Baseline Comparisons

As shown in Table 2, baseline characteristics of behavioral outcomes between intervention and control groups revealed several significant differences. Children in the control group were more likely to walk to school than those in the treatment group (p = 0.0130). Differences were also found in screen-related behaviors—such as screen use for teaching (p = 0.0884), calming (p = 0.0763), and avoiding disturbance (p = 0.0780)—as well as in mothers’ daily screen time (p = 0.009). These variations indicate a potential violation of the parallel trends assumption for these variables, limiting the strength of causal inference from Difference-in-Differences (DID) analyses for them.

**Table 2:**
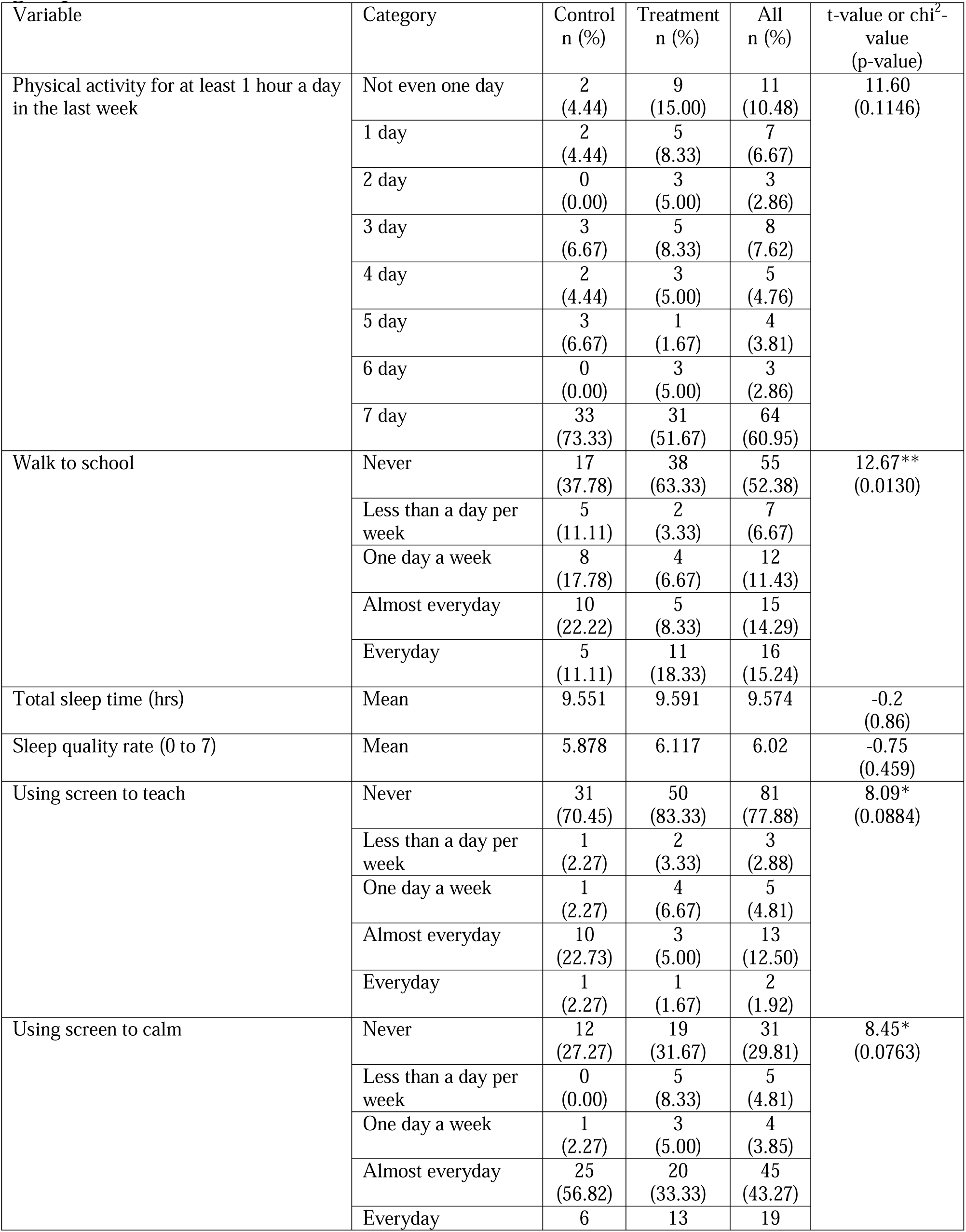
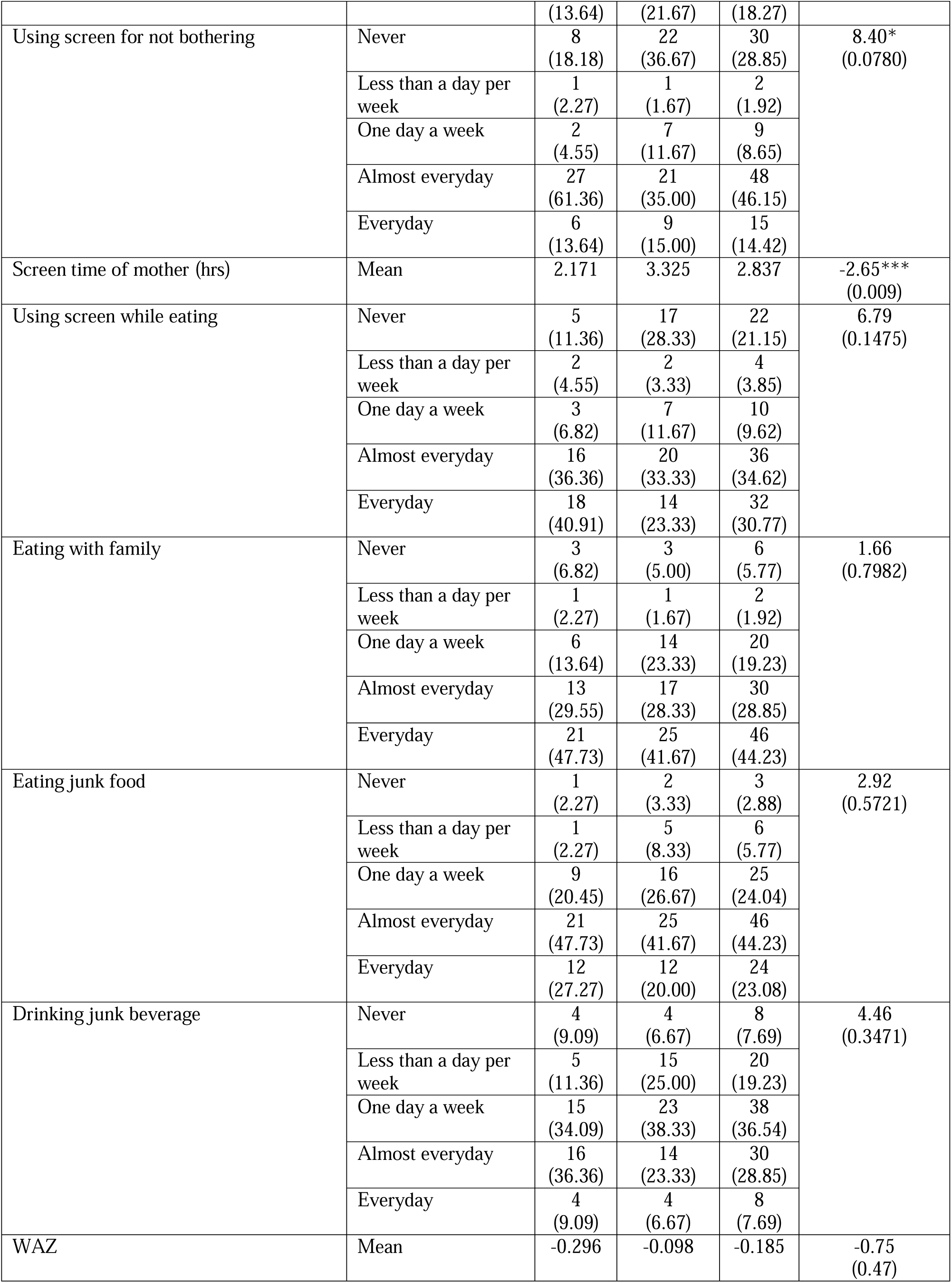
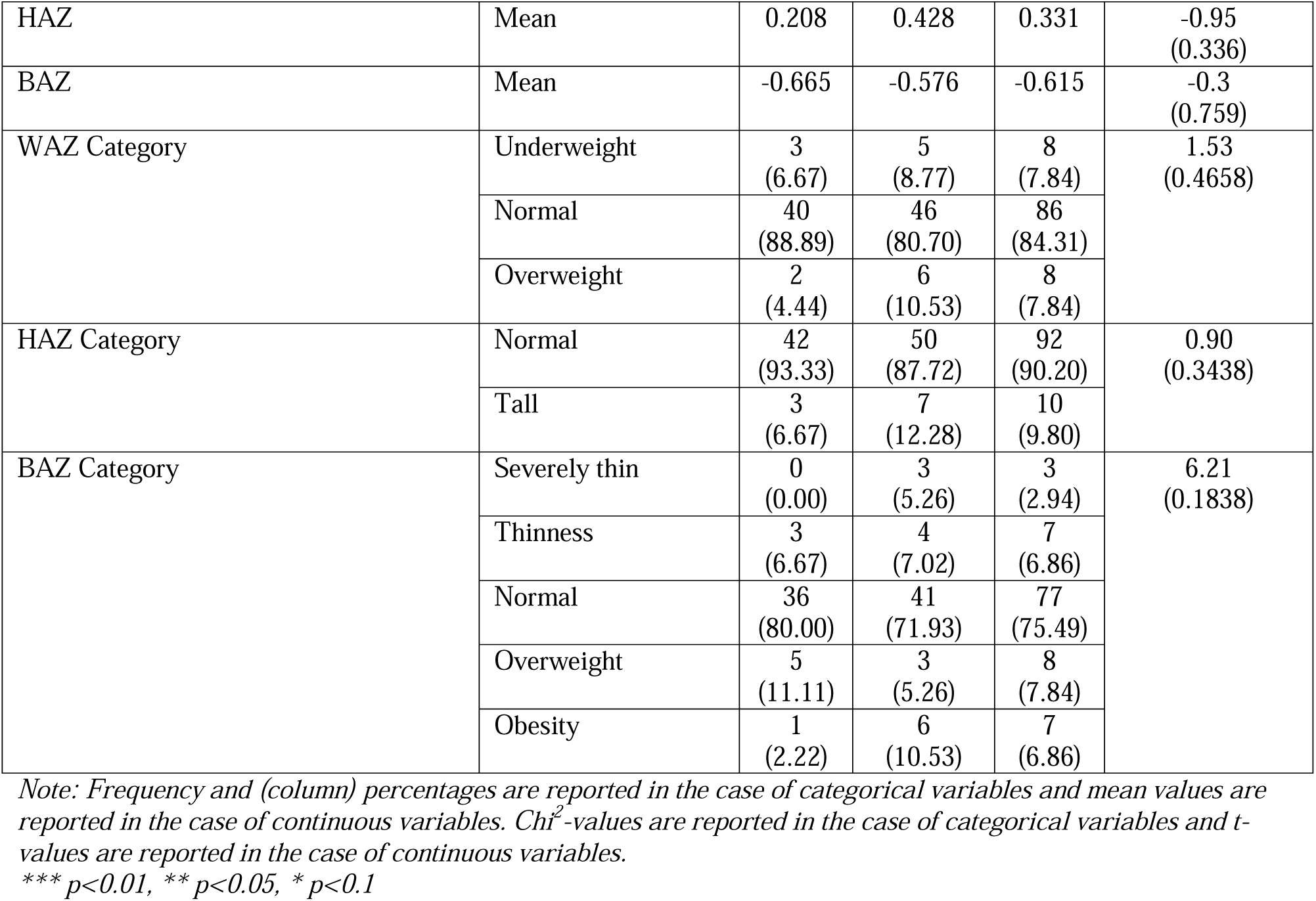
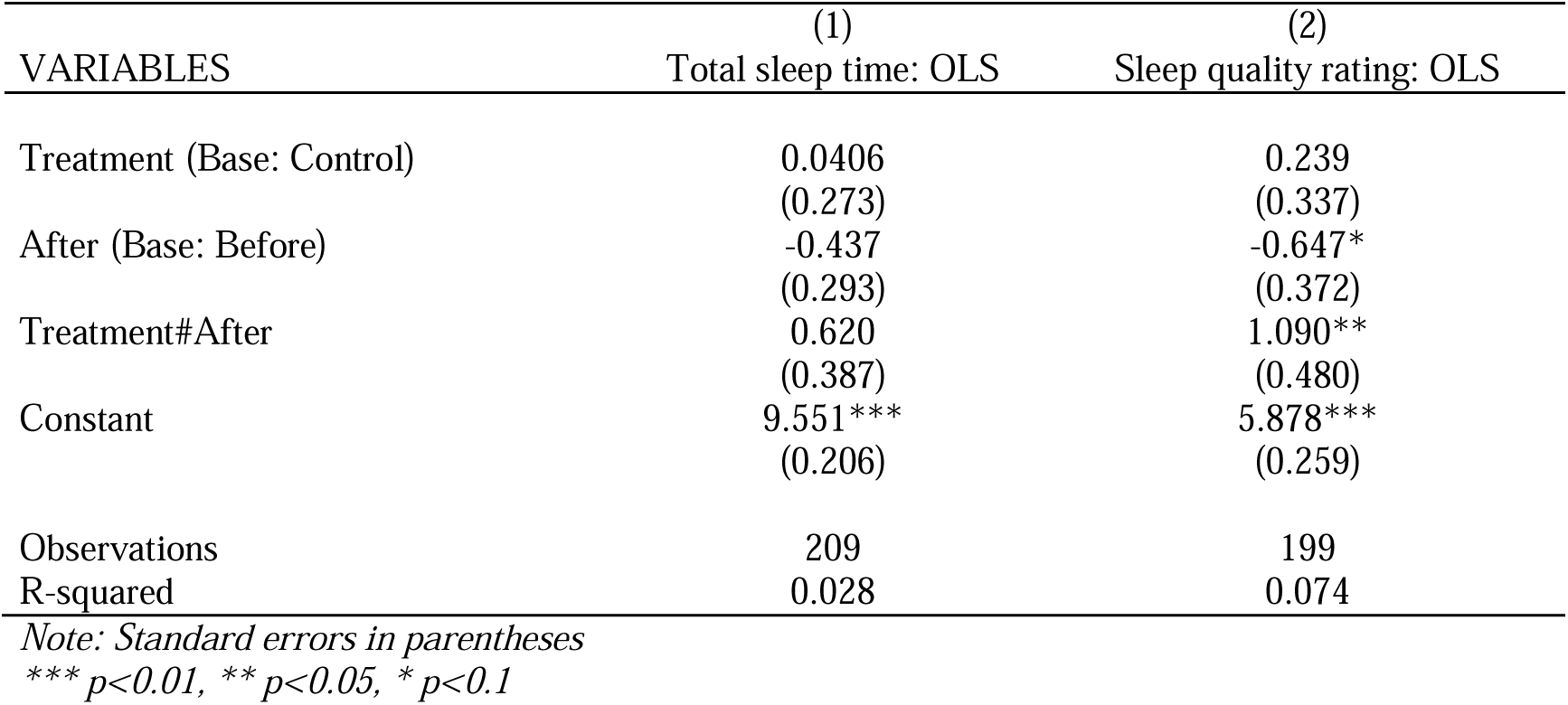
Comparison of baseline characteristics (pre-intervention) for control vs treatment group.

### Behavioral Outcomes

Pre-post comparisons (shown in Table 3) demonstrated positive behavioral changes in the intervention group. A significant increase in physical activitye was observed in the number of children engaging in ≥1 hour of physical activity per day in the intervention group (p = 0.0937), while no meaningful change occurred in the control group. The DID regression confirmed a significant intervention effect on physical activity frequency (Table 4a, β = 0.661, p < 0.05).

**Table 3:**
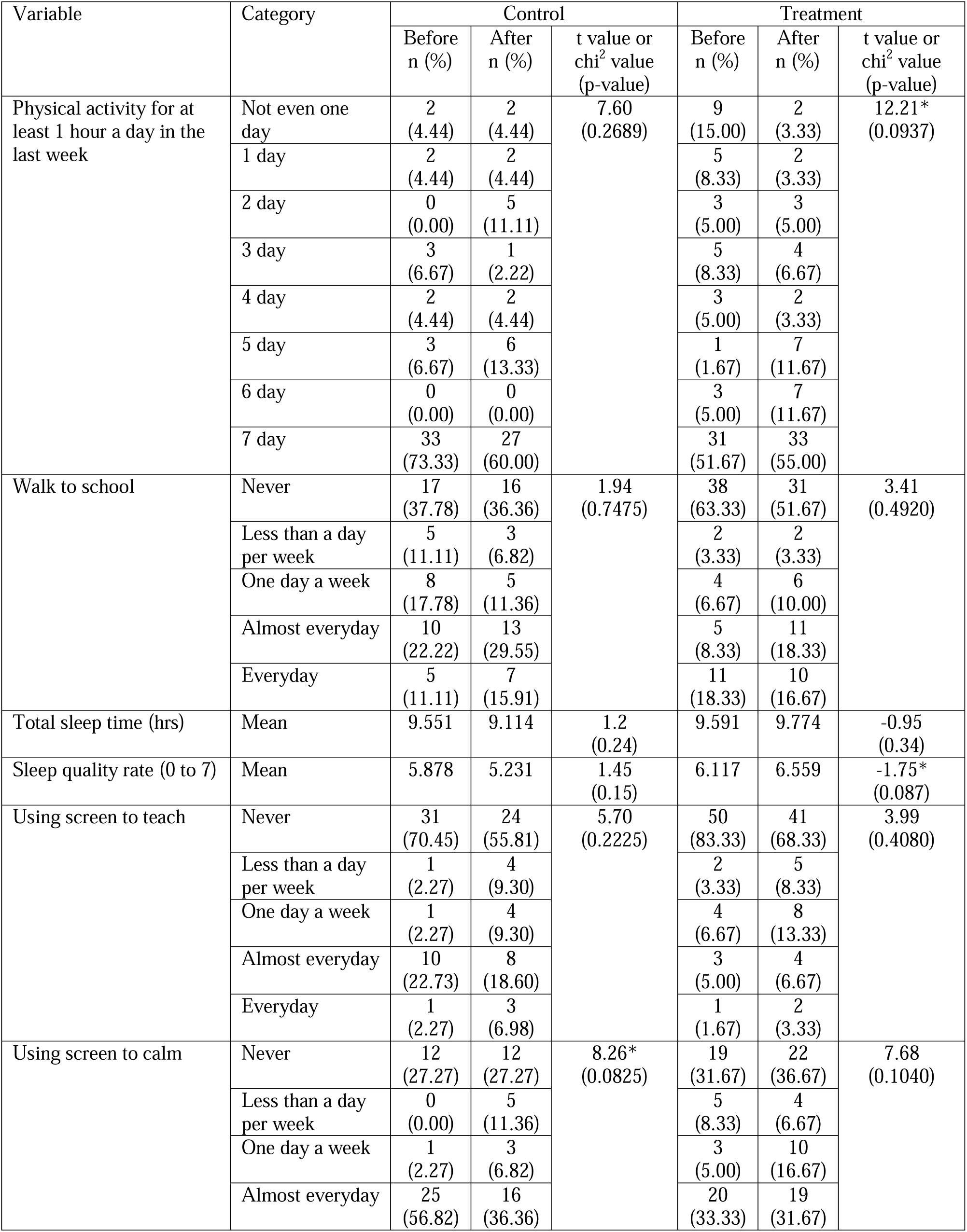
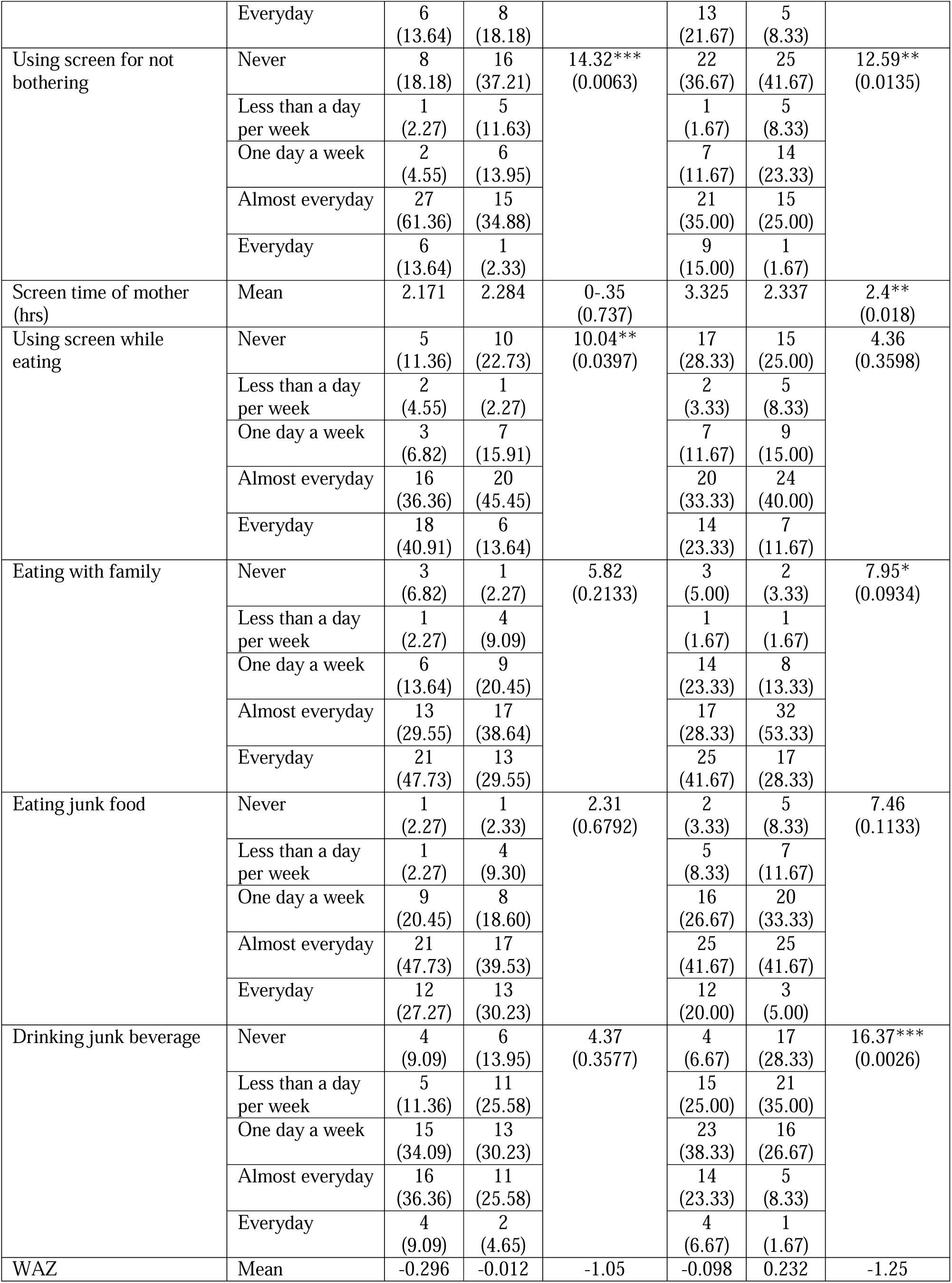
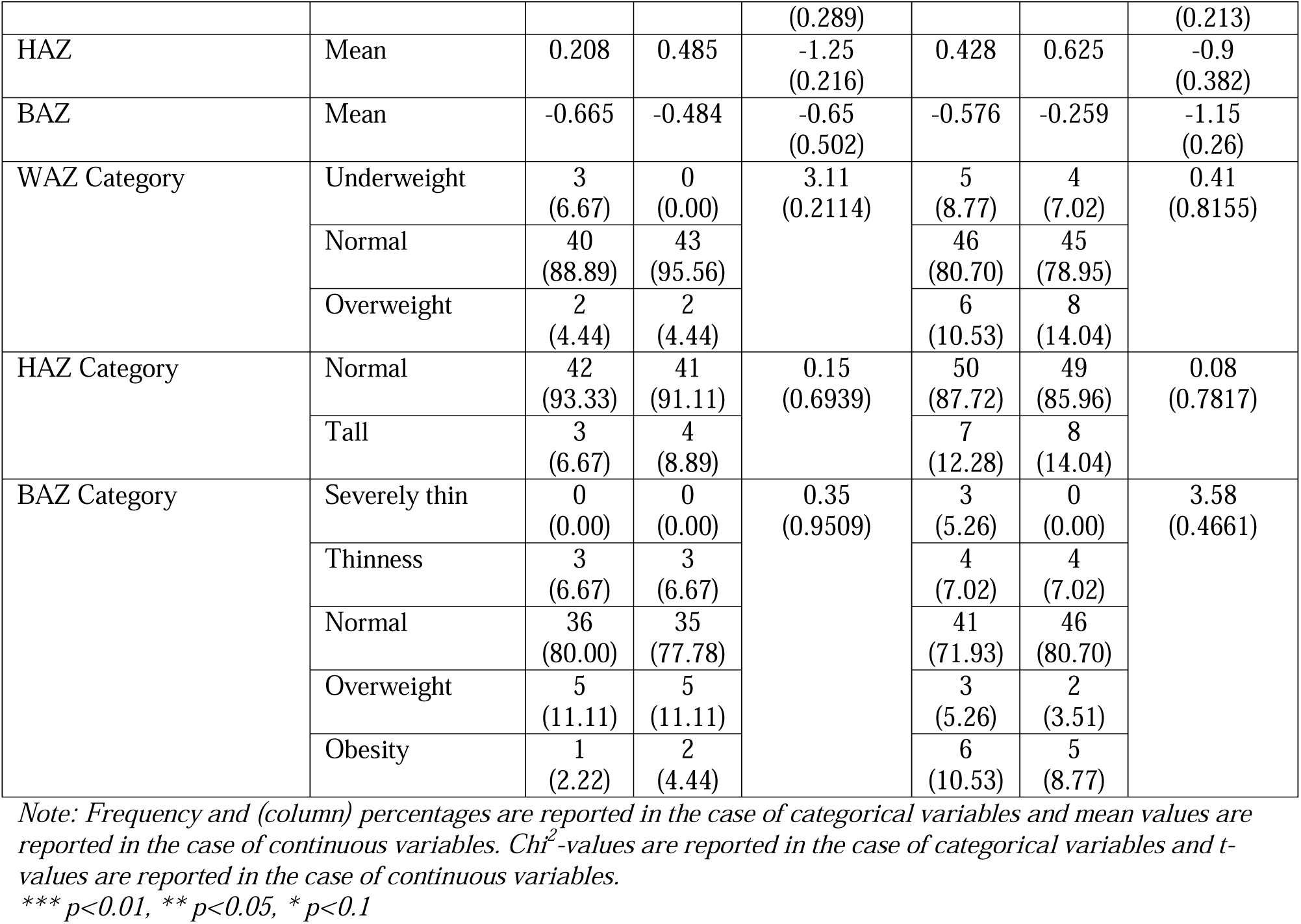
Before and after comparison for both the control group and the treatment group.

**Table 4a:** Difference-in-difference regression results: Impact on physical activity.

| VARIABLES | (1)<br>Physical activity for at least 1<br>hour a day: OP | (2)<br>Walk to school: OP |
| --- | --- | --- |
| Treatment (Base: Control) | -0.646***<br>(0.239) | -0.301<br>(0.224) |
| After (Base: Before) | -0.308<br>(0.257) | 0.140<br>(0.231) |
| Treatment#After | 0.661**<br>(0.331) | 0.0651<br>(0.314) |
| Constant cut5 | -0.924***<br>(0.194) |  |
| Constant cut6 | -0.693***<br>(0.192) |  |
| Constant cut7 | -0.566***<br>(0.191) |  |
| Constant cut1 | -1.841***<br>(0.219) | -0.109<br>(0.167) |
| Constant cut2 | -1.520***<br>(0.207) | 0.0384<br>(0.167) |
| Constant cut3 | -1.285***<br>(0.201) | 0.330**<br>(0.168) |
| Constant cut4 | -1.060***<br>(0.196) | 0.933***<br>(0.178) |
| Observations | 210 | 209 |
| Pseudo R-squared | 0.013 | 0.007 |

*Note: Standard errors in parentheses*
\*\*\* $p < 0.01$ , \*\* $p < 0.05$ , \* $p < 0.1$

Sleep quality improved significantly in the intervention group (p = 0.087) but not in the control group. DID analysis showed a positive and significant effect on sleep quality (Table 4b, β = 1.090, p < 0.05), although no significant effect was detected on total sleep time.

**Table 4b:**
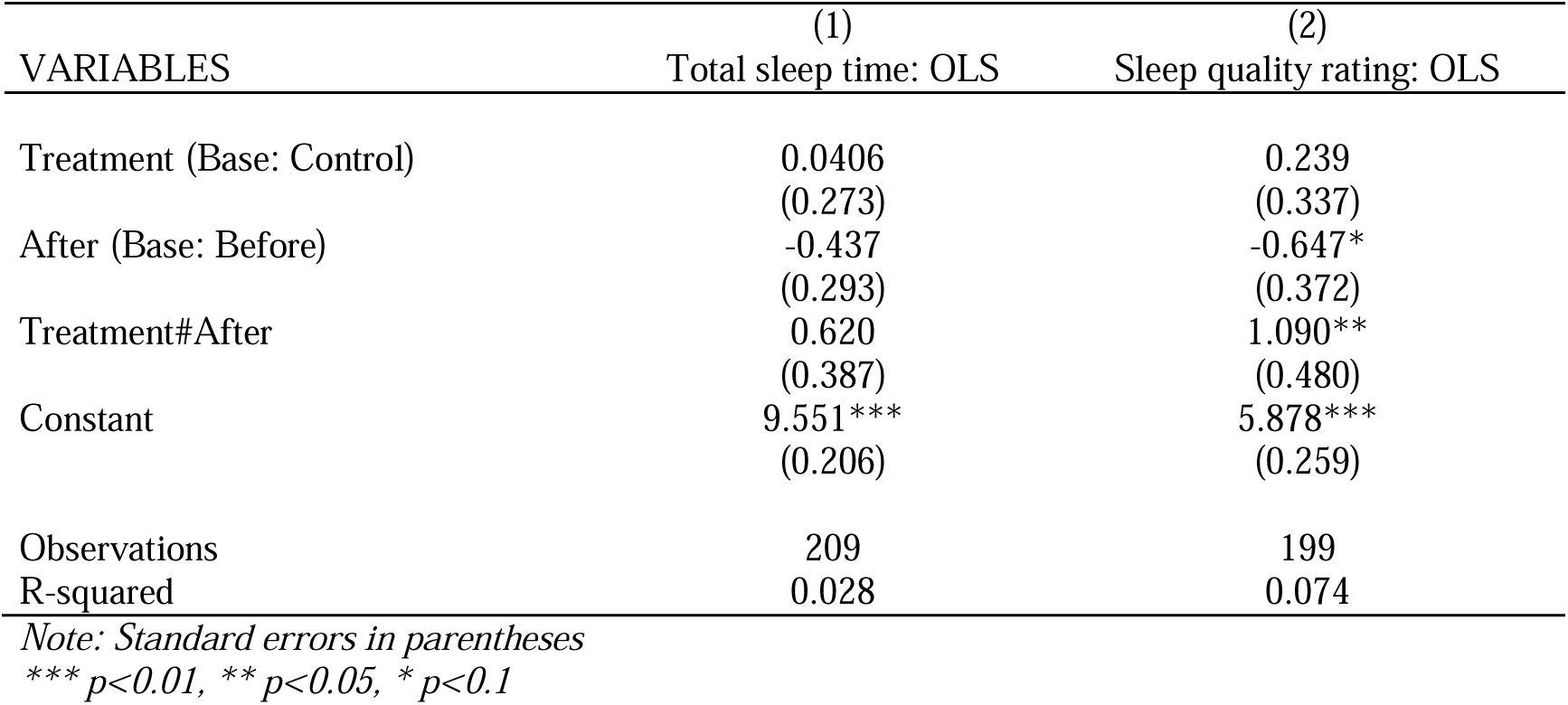
Difference-in-difference regression results: Impact on sleep.

Mothers in the intervention group significantly reduced their daily screen time (β = –1.102, p < 0.1; Table 4c, model 4). However, no significant changes were observed in children’s screen use patterns, including use for teaching, calming, or distraction purposes.

**Table 4c:**
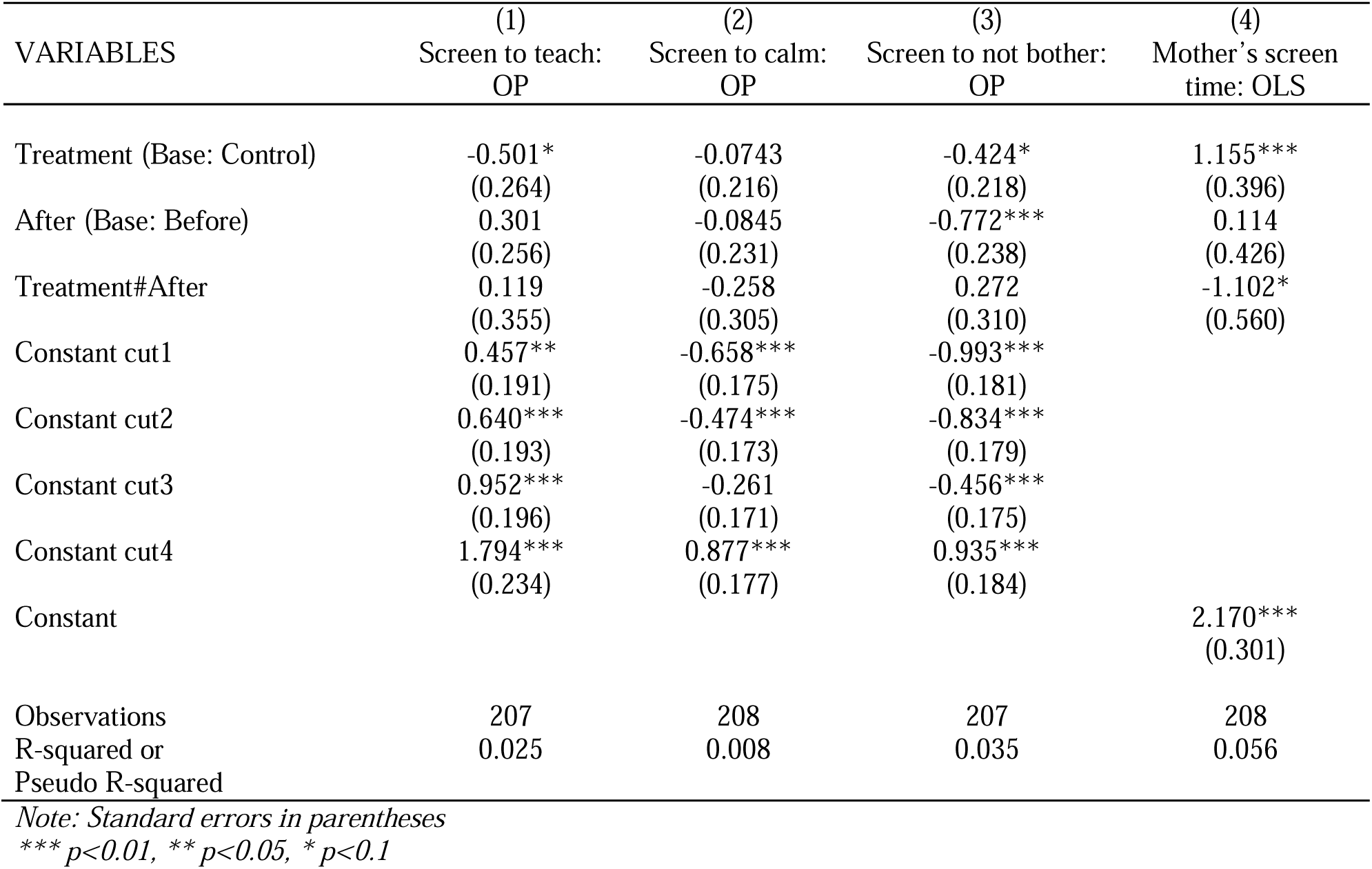
Difference-in-difference regression results: Impact on screen time.

### Dietary Behaviors

As illustrated in Table 4d, no statistically significant treatment effects were found in dietary habits. Variables such as eating with family, screen use while eating, consumption of junk food, and sugary beverage intake did not show significant changes between or within groups over time, though some descriptive trends were observed.

**Table 4d:** Difference-in-difference regression results: Impact on food habit.

| VARIABLES | (1)<br>Screen while<br>eating: OP | (2)<br>Eating with<br>family: OP | (3)<br>Eating junk<br>food: OP | (4)<br>Drinking junk<br>beverage: OP |
| --- | --- | --- | --- | --- |
| Treatment (Base: Control) | -0.588***<br>(0.218) | -0.135<br>(0.220) | -0.321<br>(0.214) | -0.260<br>(0.208) |
| After (Base: Before) | -0.631***<br>(0.233) | -0.331<br>(0.233) | -0.0802<br>(0.231) | -0.429*<br>(0.226) |
| Treatment#After | 0.463<br>(0.304) | 0.285<br>(0.306) | -0.387<br>(0.302) | -0.358<br>(0.297) |
| Constant cut1 | -1.303***<br>(0.185) | -1.882***<br>(0.215) | -2.124***<br>(0.223) | -1.589***<br>(0.189) |
| Constant cut2 | -1.149***<br>(0.182) | -1.596***<br>(0.199) | -1.538***<br>(0.192) | -0.738***<br>(0.173) |
| Constant cut3 | -0.795***<br>(0.178) | -0.827***<br>(0.179) | -0.657***<br>(0.173) | 0.182<br>(0.166) |
| Constant cut4 | 0.291*<br>(0.172) | 0.184<br>(0.173) | 0.571***<br>(0.172) | 1.260***<br>(0.195) |
| Observations | 208 | 208 | 207 | 207 |
| Pseudo R-squared | 0.022 | 0.004 | 0.030 | 0.045 |

*Note: Standard errors in parentheses*
\*\*\* $p < 0.01$ , \*\* $p < 0.05$ , \* $p < 0.1$

### Anthropometric Measures

The intervention did not lead to significant changes in weight-for-age (WAZ), height-for-age (HAZ), or BMI-for-age (BAZ) z-score categories. DID models presented in Table 4e confirmed the absence of statistically significant treatment effects on any anthropometric outcome.

**Table 4e:** Difference-in-difference regression results: Impact on anthropometry.

| VARIABLES | (1)<br>WAZ category: OP | (2)<br>HAZ category: OP | (3)<br>BAZ category: OP |
| --- | --- | --- | --- |
| Treatment (Base: Control) | 0.146<br>(0.277) | 0.340<br>(0.358) | -0.0673<br>(0.244) |
| After (Base: Before) | 0.260<br>(0.301) | 0.153<br>(0.390) | 0.0668<br>(0.258) |
| Treatment#After | -0.0767<br>(0.393) | -0.0711<br>(0.490) | 0.0367<br>(0.345) |
| Constant cut1 | -1.385***<br>(0.225) | 1.501***<br>(0.288) | -2.182***<br>(0.283) |
| Constant cut2 | 1.553***<br>(0.233) |  | -1.379***<br>(0.205) |
| Constant cut3 |  |  | 1.078***<br>(0.194) |
| Constant cut4 |  |  | 1.492***<br>(0.211) |
| Observations | 204 | 204 | 204 |
| Pseudo R-squared | 0.008 | 0.013 | 0.001 |

*Note: Standard errors in parentheses*
\*\*\* $p < 0.01$ , \*\* $p < 0.05$ , \* $p < 0.1$

## Discussion

This study aimed to evaluate the effects of a mother-centric Motivational Interviewing (MI) intervention on early childhood obesity prevention by targeting behavioral changes among preschool-aged children in a resource-limited Bangladeshi setting. While the intervention led to significant improvements in certain lifestyle behaviors—such as increased physical activity, better sleep quality, and reduced screen time among mothers—it did not result in statistically significant changes in children’s anthropometric measures. These findings provide important insights into both the potential and the limitations of community-based behavioral interventions in low- and middle-income country (LMIC) contexts.

### Behavioral Changes

The intervention successfully promoted an increase in physical activity among children, particularly achieving at least one hour of activity per day. This finding aligns with global evidence that highlights physical activity as a modifiable risk factor in childhood obesity prevention [6,8] However, walking to school did not show a significant change, possibly due to structural or environmental barriers (e.g., safety concerns, distance to school), which were beyond the scope of the intervention.

Sleep quality also improved significantly among children in the intervention group, a notable finding given the emerging literature on the importance of sufficient and good-quality sleep in regulating metabolism and preventing unhealthy weight gain[9]. Nevertheless, no significant change was observed in sleep duration, suggesting that while subjective improvements in sleep quality occurred, structural constraints such as household routines or screen time may still influence sleep patterns.

While the intervention significantly reduced mothers’ screen time—a proxy for improved awareness and modeling of healthy behaviors—it did not have a statistically significant impact on children’s screen use. Cultural norms around screen use for educational or calming purposes, and the absence of supportive structural boundaries within homes, may have diluted the intervention’s impact on children’s screen behaviors.

### Weight Status and Anthropometric Outcomes

Despite behavioral improvements, no significant changes were found in anthropometric indicators such as weight-for-age (WAZ), height-for-age (HAZ), or BMI-for-age (BAZ). One key reason for this lack of measurable change is likely the limited sample size and the short duration of the intervention, which may not be sufficient to produce detectable changes in weight trajectories in young children. Additionally, behavioral shifts—especially those related to diet and physical activity—often require sustained efforts and longer follow-up periods to influence growth patterns and adiposity. A major challenge in this study was the high dropout rate, with nearly 50% attrition, which further reduced the statistical power to detect significant differences, particularly for weight-related outcomes. High dropout not only limits the ability to generalize the findings but also introduces potential bias in estimating intervention effects, especially if attrition was non-random. Combined with the small sample size, this limits the robustness of our conclusions and may partially explain why several outcome measures—particularly those related to diet and anthropometry—did not reach statistical significance.

### Cultural Context and Parental Engagement

In developing countries like Bangladesh, childhood obesity remains a significantly under-researched area. Consequently, researchers from high-income countries often have limited awareness of the contextual challenges and realities in such settings. The *Double-Duty Action Plan*, which seeks to address both undernutrition and overnutrition, highlights school feeding as a key intervention, as reflected in the *Lancet*’s global policy recommendations. Educational institutions are identified among the top ten priorities within the plan, with particular emphasis on school feeding programs (SFPs)[20]. However, in South Asia, existing SFPs are limited in both reach and scope. Where such programs do exist, they are primarily targeted at poverty-prone regions, aiming to reduce school dropout rates and combat undernutrition. Unlike in many high-income countries, comprehensive healthy school food policies are virtually absent in this region[21].

Our study, conducted in a resource-limited, community-based setting, offers insights that could inform the design of future interventions. The study site reflected a broader cultural reality in which most mothers did not perceive childhood obesity as a health concern. Awareness of obesity-related lifestyle factors was low, and there was limited understanding of the long-term risks of obesity, including chronic non-communicable diseases.

In this context, a mother-centric MI approach—delivered in collaboration with schools— emerges as a potentially cost-effective and impactful strategy for promoting healthy lifestyle changes and preventing obesity in early childhood. The pilot study was deliberately designed to avoid disrupting regular school activities. Nevertheless, we encountered several challenges in implementing the motivational counseling sessions. Greater integration of MI into the school system could enhance its acceptability and effectiveness.

In South Asian cultures, parents are often deeply invested in their children’s academic success. Previous research has indicated a link between childhood obesity and academic performance. Incorporating such evidence into MI sessions may help reinforce parental engagement and support behavior change.

Building on lessons learned from this pilot, we have recently introduced a school diary–based lifestyle monitoring system, focusing on physical activity, screen time, sleep, and dietary habits. Parents are required to complete the diary biweekly, and it has been adopted by school authorities as part of regular practice. Preliminary observations suggest that this system is a promising tool for supporting behavior change and monitoring children’s lifestyle patterns effectively.

### Limitations

In addition to the small sample size and high attrition, another limitation was the violation of the parallel trends assumption for some baseline variables, which may compromise the internal validity of the Difference-in-Differences (DID) analysis for certain outcomes. Furthermore, the reliance on self-reported measures (physical activity, screen time, sleep) may introduce social desirability or recall biases.

## Conclusion

This pilot study demonstrates that a mother-centric Motivational Interviewing approach can effectively promote positive behavioral changes—such as increased physical activity, improved sleep quality, and reduced screen time among mothers—in a resource-limited community setting in Bangladesh. These early lifestyle modifications are essential components of childhood obesity prevention. However, the intervention did not yield statistically significant improvements in children’s anthropometric outcomes, likely due to the short intervention period, small sample size, and high dropout rate.

Despite these limitations, the findings underscore the potential of culturally tailored, school-linked MI interventions in LMICs, particularly when directed at highly engaged caregivers such as mothers. The integration of supportive tools, like school diary-based monitoring systems, offers a promising avenue for sustaining healthy behaviors. Future interventions should aim to strengthen retention strategies, extend follow-up periods, and incorporate multi-level supports— including school and policy-based components—to achieve more substantial and lasting impacts on childhood weight status.

This study provides important baseline work for designing scalable, context-sensitive obesity prevention strategies in similar low-resource settings.

## Data Availability

All data produced in the present work are contained in the manuscript

## List of abbreviations

BDT: Bangladeshi Taka;
REDCap: Research Electronic Data Capture;
SD: Standard Deviation;
DD: Difference-in-Differences

## Declarations

### Ethics approval and consent to participate

The study protocol was approved by the institutional review board of the Independent University, Bangladesh (Ref. no: 2022-SELS-02). Written informed constent was taken from earch participating mother.

### Consent for publication

Not applicable.

### Availability of data and materials

The datasets used and/or analyzed during the current study are available from the corresponding author upon reasonable request.

### Authors’ contributions

MSH, SF and ADO conceived idea and designed the study. MSH and SF were involved in data generation. FI analyzed data. MSH and FI prepared the first draft of the manuscript. All authors read, provided critical feedback and approved the manuscript.

## Acknowledgements

We would like to acknowledge SM Abdullah Al Mamun, Khondoker Amzad Hussain ebng Zubair. Zajbe (Biomedical Research Foundation, Bangladesh) for supporting data collection.

